# Multi-strain probiotic enhances metformin tolerance by modulating gut microbiome and bile acid pathways: Insight from multi-omics post-hoc analysis (ProGasMet trial)

**DOI:** 10.64898/2026.02.06.26345743

**Authors:** Hanna Kwiendacz, Danuta Cembrowska-Lech, Karolina Skonieczna-Żydecka, Karolina Klimontowicz, Konrad Podsiadło, Anna Wierzbicka-Woś, Daniel Styburski, Mariusz Kaczmarczyk, Janusz Gumprecht, Igor Łoniewski, Katarzyna Nabrdalik

**Author notes:** Contributed equally as a first-author (Hanna Kwiendacz, Danuta Cembrowska-Lech). Contributed equally as a senior-author (Igor Łoniewski, Katarzyna Nabrdalik). Corresponding author: Igor Łoniewski.

## Abstract

**Background:** Metformin is the cornerstone therapy for type 2 diabetes, but gastrointestinal intolerance commonly limits dose escalation and long-term adherence. In the ProGasMet trial, multi-strain probiotic supplementation improved metformin tolerability. However, the underlying microbiome-metabolome mechanisms remain unclear.

**Methods and analysis:** We performed an exploratory multi-omics analysis using Period 1 of a randomized, double-blind, placebo-controlled trial. Participants with metformin intolerance received a multi-strain probiotic or placebo for 12 weeks. Paired stool samples collected at baseline (Visit 2) and end of treatment (Visit 5) were available from 34 participants (68 samples). We integrated shotgun metagenomic species profiles, predicted gut metabolic modules, and untargeted faecal LC-MS metabolomics using multi-block sparse PLS (DIABLO), complemented by longitudinal feature-level analyses and associations with gastrointestinal symptom burden (QACSMI and a simplified GI score).

**Results:** Multi-omics integration showed moderate concordance across taxonomic, functional, and metabolomic blocks and separated probiotic from placebo profiles at 12 weeks. Bile acid-related metabolites were among the strongest contributors to group separation, with hyodeoxycholic acid and related compounds enriched in the probiotic arm. Global biodiversity and community-wide turnover did not differ materially between groups. Feature-level analyses suggested modest, directionally coherent changes in selected taxa, functional modules, and metabolites. Higher hyodeoxycholic acid concentrations at Visit 5 were associated with lower gastrointestinal symptom burden in probiotic-treated participants, a pattern not observed under placebo; statistical support was exploratory.

**Conclusion:** Probiotic supplementation may be associated with coordinated microbiome-metabolome shifts in metformin-intolerant type 2 diabetes, highlighting bile acid remodelling, particularly hyodeoxycholic acid, as a plausible mechanistic candidate for improved tolerability.

## 1. Background

According to the American Diabetes Association (ADA) and European Association for the Study of Diabetes (EASD) consensus guidelines, metformin remains one of the most frequently used antyhiperglycemic agents in individuals with T2DM [1,2]. However, metformin use, especially in optimal doses, is in some cases limited by gastrointestinal (GI) adverse effects (AEs). The mechanisms underlying metformin-induced GI intolerance are incompletely understood and appear multifactorial. Growing evidence indicates that the gut microbiota is a major mediator of both metformin efficacy and intolerance. Metformin is now recognized as one of the most influential non-antibiotic drugs acting on gut microbiota composition and function [3–5]. Experimental work shows that intravenous metformin has little effect on glucose homeostasis, whereas oral administration exerts robust antihyperglycemic effects, suggesting that GI passage and local gut actions are critical [6,7]. Fecal microbiota transplantation from metformin-treated donors to germ-free mice improves glucose tolerance, and antibiotic-mediated depletion of the microbiota abrogates metformin’s metabolic benefits in animal models, underscoring the modulatory role of the gut microbiome in mediating metformin’s actions [7–9]. In humans, metformin treatment has been repeatedly associated with shifts in the gut microbiome, including increased abundance of taxa such as *Akkermansia* and *Escherichia*-*Shigella* and reduced levels of Intestinibacter and several butyrate-producing genera [3,10–12]. Some of these changes, particularly enrichment of opportunistic pathobionts and short-chain fatty acid (SCFA)-producers, may contribute at once to improvement in glycaemic control and to GI symptoms [13–15].

Beyond taxonomic shifts, T2DM itself is associated with an altered gut ecosystem characterised by reduced diversity and depletion of butyrate-producing bacteria such as *Faecalibacterium prausnitzii*, with consequences for intestinal barrier integrity, endotoxemia and low-grade inflammation [16–21]. Diet and geography further modulate microbiota composition, suggesting that microbiome-derived metabolites may be more conserved biomarkers of metabolic status than individual taxa [20,22,23]. These metabolites can provide mechanistic insights and may serve as targets for novel therapeutic strategies, particularly in the context of impaired glucose metabolism.

Bile acids sit at a key interface between the gut microbiome, host metabolism and metformin’s actions. In addition to facilitating lipid absorption, they act as signaling molecules that regulate their own synthesis and glucose and lipid homeostasis via the Farnesoid X Receptors (FXR) and Takeda G protein-coupled receptor 5 (TGR5) [24–28]. Metformin reduces ileal bile acid reabsorption and alters intestinal bile acid exposure, which may contribute both to its glucose-lowering effect and to gastrointestinal symptoms [14,28,29]. Because gut bacteria deconjugate and transform primary into secondary bile acids, and probiotic strains can modify these reactions [30–32], bile acid remodeling represents a plausible pathway through which microbiota-targeted interventions might influence metformin tolerance.

In contrast to this mechanistic backdrop, microbiota modulation has emerged as a promising strategy for reducing metformin-associated GI intolerance. Probiotics are among the best-studied microbiome modulators. They may restore microbial balance, enhance epithelial barrier function, influence bile acid metabolism and tune immune and enteroendocrine responses [32–34]. Clinical evidence in metformin-intolerant patients is accumulating. A meta-analysis of randomized controlled trials suggested that probiotic supplementation can reduce GI discomfort associated with metformin and may modestly improve glycaemic control, although heterogeneity and risk of bias limit firm conclusions [35,36]. In the ProGasMet trial, a 32-week randomised, double-blind, placebo-controlled, cross-over study, we previously showed that a multi-strain probiotic (Sanprobi Barrier) significantly reduced the incidence, frequency and severity of nausea and abdominal bloating/pain as well as improved self-assessed metformin tolerability in individuals with T2DM and metformin intolerance [15]. Similar symptomatic benefits in metformin-treated T2DM individuals have been reported for other probiotic formulations, such as *Bifidobacterium bifidum* G9-1, who presented with GI symptoms without altering HbA_1c_, suggesting that symptoms relief can be achieved independently of glycaemic changes [37].

However, important gaps in knowledge remain. Most existing studies, including our own clinical ProGasMet report, have focused primarily on clinical outcomes, without establishing direct mechanistic links between specific microbial taxa, functional pathways, metabolites and reduced metformin intolerance. The microbiome– and metabolome-related characteristics underpinning the beneficial clinical effect observed in our previous report of ProGasMet study [15], however, remained unclear. The present paper is a continuation of ProGasMet data analysis, which addresses this gap by applying an integrative exploratory multi-omics approach encompassing gut microbiome taxonomic composition (shotgun metagenomics), predicted metabolic modules, and untargeted faecal metabolomics, together with complementary univariate analyses. Our aims were to: (i) characterize multi-omic signatures associated with probiotic versus placebo supplementation in metformin-intolerant T2DM participants using an exploratory integrative analysis; (ii) identify specific microbial taxa, functional pathways and metabolites, to generate mechanistic hypotheses on how probiotics may mitigate metformin GI side effects.

## 2. Materials and methods

### 2.1 Study design

This is a post-hoc multi-omics analysis of ProGasMet study data (ProGasMet; ClinicalTrials.gov identifier NCT04089280) which was a 32-week, single-centre, randomized, double-blind, placebo-controlled, two-period cross-over clinical trial involving adults with T2DM who experienced GI intolerance to metformin. The inclusion as well as exclusion criteria, study population, and study procedures were described in detail previously [15]. Briefly, participants were randomly allocated in a 1:1 ratio to one of two treatment sequences via a permutation–based randomization list: multi-strain probiotic (Sanprobi Barrier, manufacturer – Sanprobi sp. z o.o. sp. k, Poland, containing *Bifidobacterium bifidum W23, Bifidobacterium lactis W51, Bifidobacterium lactis W52, Lactobacillus acidophilus W37, Levilactobacillus brevis W63, Lacticaseibacillus casei W56, Ligilactobacillus salivarius W24, Lactococcus lactis W19, and Lactococcus lactis W58* administered 2 x 2 capsules daily in the daily dose 2 x 10^9^ colony forming units, CFU), followed by placebo, or the reverse sequence, in two 12-week treatment periods separated by a 4-week washout, with ongoing metformin therapy and protocolised metformin titration.

In the present multi-omics analysis, we restricted our dataset to the first 12–week treatment period and considered only two timepoints: baseline (Visit 2, V2) and the end of the initial treatment period after 12 weeks of probiotic or placebo supplementation (Visit 5, V5), effectively treating the comparison as parallel-group probiotic versus placebo. This restriction was implemented to eliminate the influence of carry-over effect observed in the parent trial (Nabrdalik et al., 2023). The overall trial flow and clinical analysis population have been reported previously (Nabrdalik et al., 2023). Of the participants completing Period 1 and 2 (35 individuals), one participant the fecal metabolomics profile at both V2 and V5 did not yield quantifiable features after quality control and filtering (signals were below the detection threshold), resulting in 34 participants (68 samples) with complete paired multi-omics profiles for integrative analyses.

### 2.2 Outcome measures

The primary outcomes of this exploratory multi-omics study were microbiome– and metabolome-derived measures: (i) species-level taxonomic composition, (ii) microbial functional potential quantified as gut metabolic modules (GMMs), and (iii) faecal metabolite profiles. These endpoints were analysed to characterise multi-omic signatures associated with probiotic versus placebo supplementation and to identify candidate taxa, pathways and metabolites underpinning inter-individual responses.

Secondary outcomes were linking molecular characteristics with clinical tolerance to metformin and to propose mechanistic hypotheses of its GI side effects. Symptoms were quantified using the Questionnaire to Assess Character and Severity of Metformin Intolerance (QACSMI; Metformin Symptom Severity Score), adapted from [38], which captures the character, frequency and severity of metformin-associated gastrointestinal symptoms. In addition, we derived a simplified gastrointestinal symptom score (GI) as the summed burden of three core symptom domains (nausea, bloating pain and diarrhea) each recorded in terms of frequency and severity. In this post-hoc multi-omics analysis, QACSMI and GI were used only as quantitative symptom phenotypes to examine associations with microbial taxa, functional modules and metabolites.

### 2.3 Sample collection

Stool samples were collected into stool sampling containers, frozen, and stored at –80°C until further processing at time points previously described by Nabrdalik et.al. [15].

### 2.4 Fecal metagenomic sequencing

#### 2.4.1 DNA isolation and shotgun metagenomics

Metagenomic DNA was extracted using the QIAmp PowerFecal Pro DNA kit (Qiagen), following the manufacturer’s instructions, with additional mechanical disruption via bead–beating homogenization on the PowerLyzer 24 device (Qiagen). Shotgun sequencing libraries were prepared from 30 ng of extracted DNA using the KAPA HyperPlus kit (ROCHE) with a 1:3 reduced reagent volume, as described by Sanders (2019). Sample indexing was performed through ligation with KAPA Unique Dual-Indexed adaptors (ROCHE), and DNA fragments of approximately 300–400 bp were selected for further steps. The quality and quantity of DNA were assessed using spectrophotometric and fluorometric methods on the DeNovix DS–11 device (DeNovix). Double-stranded DNA (dsDNA) concentrations were measured using QuantiFluor dsDNA ONE dye (Promega). Library quality and fragment lengths were evaluated for NGS suitability using microchip electrophoresis on a MultiNA device (SHIMADZU) with the DNA-1000 kit (SHIMADZU). Next–generation sequencing (NGS) was performed on the Illumina MiSeq platform using a paired-end strategy (2 × 151 bp). Shallow shotgun sequencing was employed to profile the microbial community composition and the functional potential of the microbiome present in the samples.

#### 2.4.2 Microbiome data processing

To process the raw shotgun metagenomic sequencing data, the BioBakery workflows (v3.1) pipeline was employed. The quality control process began with KneadData v0.12.1, which incorporates Trimmomatic (v0.33), Bowtie2 (v2.5.4), and Python (v3.10.16). Trimmomatic was used to remove low-quality bases and adapter sequences. Bowtie2 was applied to align and eliminate host-derived reads. Additionally, TRF v4.09.1 was used to mask tandem repeats. The average number of reads per sample was 578,129 (range: 243,301–901,327).

Taxonomic and functional annotation were performed using NGLess v1.5.0 [39]. The mOTU module (version 3.1) was used to obtain the taxonomic profiles from the mOTU to phylum level in absolute abundance (raw counts, relative_abundance=False) using the default alignment length of 75 nucleotides. For functional annotation, reads were mapped against the Integrated Gene Catalog (IGC) [40], a comprehensive human gut microbiome database using the minimum match size of 90 nucleotides and 90% minimum identity. KEGG Orthologs (KO) counts were obtained without feature–length normalization (normalization={raw}) and all mapping hits were counted separately (multiple={all1}). Gut Metabolic Modules (GMMs) [41] were computed from KO abundance matrix using the omixer-rpmR library (version 0.3.3) using a minimum coverage = 0.3. After KO-to-GMM mapping, library-size normalization was applied for cross–sample comparability. Alpha-diversity metrics (richness, evenness, Shannon, and Simpson) were calculated from a rarefied species count table (rarefaction depth = 120). Rarefaction was performed using the rtk R library (version 0.2.6.1), and alpha-diversity values were computed using the get.mean.diversity function. Beta-diversity was assessed using Bray-Curtis dissimilarities calculated from the same rarefied species data with the vegan R package (version 2.7-2). To compare beta-diversity between intervention groups, within-subject Bray-Curtis distances were extracted from the full distance matrix.

Raw taxonomic count data were processed using the ALDEx2 package (version 1.36.0) in R [42], which applies a centered log–ratio (CLR) transformation with Monte Carlo sampling from the Dirichlet distribution to account for compositional data. CLR values were generated using the aldex.clr function with 1000 Monte Carlo instances and the interquartile log–ratio denominator (denom = “iqlr”), which computes the geometric mean from features with typical between–sample variance to obtain a more stable CLR normalization. For a robust denominator estimate, the CLR transformation was first applied to the complete dataset, and feature filtering was performed after CLR computation. Features were retained if they were detected in at least 10 % of samples (count > 0 in the raw taxonomic count data). For each feature and sample, the median CLR value across Monte Carlo instances was used to obtain a single abundance matrix for downstream analyses.

### 2.5 Fecal untargeted metabolomics

#### 2.5.1 Sample preparation and untargeted metabolome profiling

To prepare the samples, was added 500 µl of a mixture containing methanol, water, and acetonitrile (in a 50:25:25, v/v/v ratio) along with deuterated internal standards to 60 mg of feces. The mixture was then shaken at 2000 rpm at 4°C for 30 minutes to dissolve metabolites and precipitate proteins. Next, the samples were centrifuged at 4000 rpm for 4 minutes at 4°C. After centrifugation, the supernatant was filtered through a 0.22 μm syringe filter into chromatography tubes. All samples were analyzed on the same day using liquid chromatography–mass spectrometry (LC-MS).

The analysis was performed using an ExionLC liquid chromatograph equipped with a binary pump, autosampler, and column thermostat connected to a Triple TOF 6600+ mass spectrometer (Sciex, Framingham, MA, USA). We used a Phenomenex Luna® Omega 1.6 μm polar C18 150 x 2.1mm column for separation, which took about 45 minutes using a gradient method. The mobile phases consisted of Phase A (water with 10mM ammonium acetate) and Phase B (acetonitrile with 0.1% formic acid). We injected 2μl of each sample, maintaining the column at 20°C with a flow rate of 0.2 ml/min. Spectral data were collected in both positive and negative ion modes. In positive mode, the capillary voltage was set to 5500 V, with a Curtain gas (CUR) of 25 psi, Ion source gas 1 (GS1) at 45 psi, Ion source gas 2 (GS2) at 60 psi, and the ion source temperature at 400°C. For negative ion mode, the capillary voltage was 4500 V, with the same gas settings, but the ion source temperature was reduced to 350°C. Spectral data were collected in DDA (Data-Dependent Acquisition) mode. This is a tandem mass spectrometry (MS/MS) mode in which the instrument automatically selects the most intense precursor ions from a full MS scan for subsequent fragmentation and analysis. This allows for the generation of fragmentation spectra for the most abundant ions. In our analysis, we selected the 10 most intense ions at a given time for fragmentation and analysis.

#### 2.5.2 Chromatographic data pre-processing and metabolite identification

Data files generated on the ExionLC system with TripleTOF 6600+ mass spectrometer (SCIEX, Framingham, MA, USA) were converted with the Reifycs Analysis Base File Converter (References: Reifycs Abf Converter. Available online: https://www.reifycs.com/AbfConverter/) and imported into MS–DIAL 4 [43] for untargeted processing. Within MS–DIAL, raw data underwent automated peak detection, MS/MS deconvolution using the MS2Dec algorithm, and retention time alignment across all samples to assemble consensus features. The deconvolution step generated purified product–ion spectra for each precursor, thereby improving spectral specificity prior to library matching, a workflow recommended for SWATH/DIA–type datasets acquired on TripleTOF instruments. Library interrogation was performed against high–resolution MS/MS resources, including HR-MS/MS, NIST, MassBank [44] and MassBank-EU (MassBank-EU. Available online: MassBank Europe. https://massbank.eu/), GNPS [45], and HMDB [46], using the NIST MSP text format to ensure broad spectral coverage and compatibility. Putative structures for the library–matched and unmatched features were further evaluated by *in silico* formula and structure elucidation with MS–Finder 3.52 [47] and SIRIUS 4.9 [48]. Final structural depictions and identifiers, including InChI Keys and canonical SMILES, were generated in ChemDraw 20.1.1 (PerkinElmer Informatics, Rodgau, Germany) to provide unambiguous chemical annotations suitable for downstream reporting.

Metabolomics data were preprocessed and normalized in Python v3.13 (PyCharm Professional v2025.2.4 IDE for Fedora Linux v41) using the packages scikit–learn v1.7.2. Raw intensity matrices were first aligned to the study metadata and filtered with a modified presence criterion (MIN_PRESENCE = 0.80). To reduce sample–to–sample loading/dilution differences, quantile normalization was then applied across samples using scikit–learn’s QuantileTransformer (output distribution set to normal), thereby enforcing a common empirical distribution of intensities. Following normalization, data were Pareto scaled at the metabolite level by means–centering each feature and dividing by the square root of its standard deviation, which stabilizes variance while preserving relative differences between metabolites and avoiding the over-equalization associated with unit–variance autoscaling.

### 2.6 Statistical and explorative analysis

#### 2.6.1 Integrative multi-omics analysis

An integrative analysis of species-level taxonomic profiles, gut metabolic modules (GMMs), and fecal metabolomics was performed using the mixOmics R package (version 6.28.0) [49]. We used the Data Integration Analysis for Biomarker discovery using Latent cOmponents (DIABLO) framework [50] to integrate these three data layers and to explain their relationship with intervention type at V5. This supervised algorithm identifies correlated features across omic layers while maximizing discrimination between predefined groups (i.e. PRO and PLA). Design matrix weights were 0.1 favouring predictive accuracy. First the model was fit without feature selection in order to assess the overall performance and optimal number of components. The performance of the fitted model was assessed using a 10–fold cross validation with 10 repeats. Based on classification error rate 3 components and centroid distances were chosen (Supplementary Fig. 1). To select the optimal number of features for each omic layer we used the tune.block.splsda function with 10–fold cross validation and 10 repeats with 3 components and centroid distances.

#### 2.6.2 Univariate analysis – primary outcomes

Alpha-diversity metrics, taxonomic features, functional modules, and gut metabolites were analysed longitudinally using general linear mixed–effects models to account for clustered data (repeated measures at V2 and V5). Mixed-effects models were fitted using the lme4 R package (version 1.1-37). Because lme4 does not provide P values for fixed effects by default, the significance of time (V2 vs. V5) by intervention group interaction was assessed using likelihood ratio tests comparing nested models with and without the interaction term. To make inferences about the relationship between the variables and to visualize them we use mode-based estimates generated using the ggeffects R package (version 2.3.1).

#### 2.6.3 Univariate analysis – secondary outcomes

To explore the potential relevance of omics features, we examined their associations with metformin-related adverse events using both QACSMI and the simplified GI score, assessed cross-sectionally at V5. General linear models (for continuous outcomes) and generalized linear models (for binary outcomes) were applied. Continuous GI outcomes were rank-transformed prior to model fitting. Binary outcomes were derived from the original (unranked) scores using median dichotomization, with QACSMI ≤ 10 and GI ≤ 7 coded as 0, and values above these thresholds coded as 1. For visualization, model-based estimates were generated using the effect R library (version 4.2-4).

All primary and secondary outcome models in univariate analysis were adjusted for age, sex, BMI, metformin dose (time-varying), and duration of diabetes mellitus.

### 2.7. Ethics

Good Clinical Practice guidelines, and the Declaration of Helsinki has been aplied to the study and the Ethics Committee of Medical University of Silesia (approval number: KNW/0022/KB1/52/I/18) provide approval for this study.

## 3. Results

### 3.1 Clinical characteristics

The characteristics of study participants (N = 34) are summarized in Table 1. The distribution of sex was comparable between groups (P = 0.296), and the mean age was similar, at 65 (± 6) years in the PRO group (i.e. probiotic group) and 62 (± 8) years in the PLA group (i.e. placebo group) (P = 0.284).

**Table 1.** Patients’ characteristics.

| Characteristic | PRO N = 17<sup>1</sup> | PLA N = 17<sup>1</sup> | P<sup>2</sup> |
| --- | --- | --- | --- |
| Sex |  |  | 0.296 |
| Female | 12 / 17 (71%) | 8 / 17 (47%) |  |
| Male | 5 / 17 (29%) | 9 / 17 (53%) |  |
| Age (years) | 65 (6) | 62 (8) | 0.284 |
| BMI, kg/m<sup>2</sup> (Visit 2) | 31.2 (5.6) | 33.4 (6.4) | 0.370 |
| BMI, kg/m<sup>2</sup> (Visit 5) | 31.5 (5.7) | 33.4 (6.0) | 0.438 |
| Metformin dose, mg (Visit 2) | 982 (460) | 1,168 (311) | 0.223 |
| Metformin dose, mg (Visit 5) | 1,688 (553) | 1,844 (560) | 0.427 |

<sup>1</sup>n / N (%), <sup>2</sup>Pearson's Chi-squared test; Kruskal-Wallis rank sum test, N (%) or mean (SD)
BMI – body mass index, PRO – probiotic, PLA – placebo, N – number, P – level of significance

At randomization (V2), the mean body mass index (BMI) was 31.2 (± 5.6) kg/m² in the PRO group and 33.4 (± 6.4) kg/m² in the PLA group (P = 0.370). By V5, BMI remained stable at 31.5 (± 5.7) and 33.4 (± 6.0) kg/m², respectively (P = 0.438). The mean daily metformin dose at V2 was 982 (± 460) mg in the PRO group and 1,168 (± 311) mg in the PLA group (P = 0.223), rising to 1,688 (± 553) mg and 1,844 (± 560) mg, respectively, by V5 (P = 0.427).

### 3.2 Integrative multi-omics exploratory analysis

The multi-omics analysis performed at V5 indicates that probiotic supplementation may induce coordinated alterations in microbial species composition, functional potential (Gut Metabolic Modules, GMMs), and gut metabolite profiles (Fig. 1). Moderate cross-omic correlations (pairwise correlation range 0.41-0.59, Fig. 1A) was observed, and sample plots confirmed a discriminative ability between the interventions (Fig. 1B). The correlation circle revealed relatively loose relationships between variables, with several features showing positive and negative correlations on both components 1 and 2 (Fig. 1C). A clustered heatmap of selected taxonomic, functional and metabolite features showed that samples at V5 segregated into two main clusters, one of which consisted almost entirely of PRO participants. Within this PRO-enriched cluster, coherent patterns emerged, including decreased levels of selected GMMs (e.g., MF0063, MF0024), increased levels of secondary bile acids and specific GMM modules, and elevated abundances of several bacterial taxa (e.g., *Clostridium* sp.; Fig. 1D). To identify the features driving the observed group separation (Fig. 1b), we performed feature importance analysis. In the species block, *Bacteroides plebeius* and *Firmicutes* sp. were more prominent in the PLA group, whereas *Clostridium* sp. and *Clostridiales* sp. contributed more strongly in the PRO group. Among GMMs, MF0024 and MF0102 were associated with the PLA group, while MF0052 and MF0072 were more characteristic of the PRO group. In the metabolomics block, bile acid-related metabolites, hyodeoxycholic acid, chenodeoxycholic acid, and 4-guanidinobutanoic acid, together with several amino acid-related compounds, showed the strongest contributions and were, on average, higher in the PRO group. In contrast, metabolites such as nutriacholic acid and oleamide were enriched in the PLA group (Supplementary Fig. 2).

**Fig. 1.**
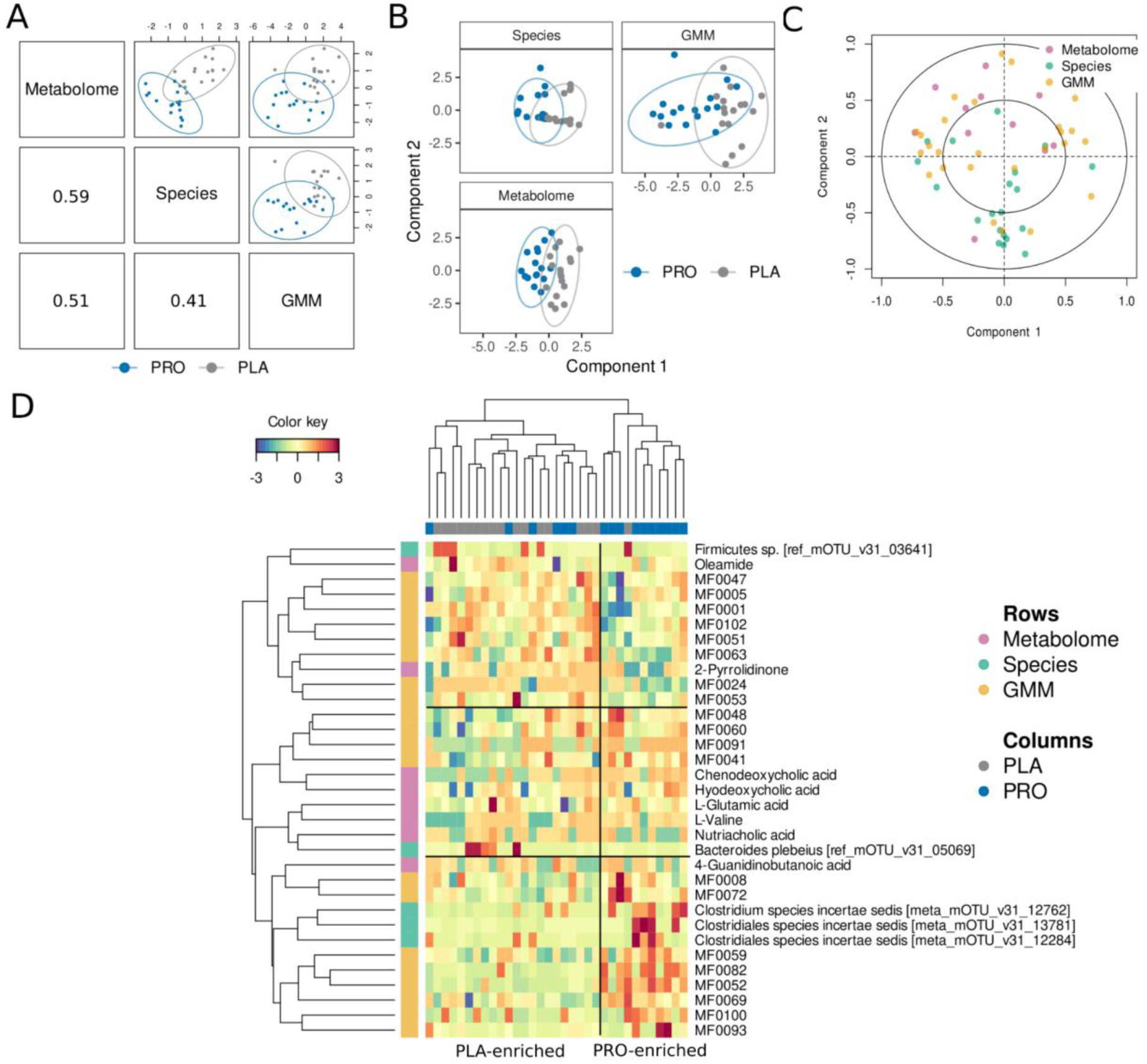
Multi-omics integration of gut microbial taxa, gut metabolic modules and stool metabolome in probiotic and placebo groups. (A) The first components extracted from the three omics datasets were moderately correlated with each other (pairwise correlation range 0.41–0.59), indicating a common structure at the taxonomic, functional, and metabolic levels. Ordination of samples along the first component of each pair of omic layers showed a separation between PRO and PLA samples, suggesting that the intervention induced a coordinated change in microbial composition, functional potential, and metabolite profiles (B) Sample plot – projections of each sample into the space spanned by the components from each block – when the first two components were examined separately for each omic dataset, probiotic and placebo samples were grouped separately (with partial overlap). (C) The correlation circle plot visualizes the contribution of each selected variable to the first two components. Variables positioned near the outer circle are those most strongly correlated with a given component. The plot displays variables from all data blocks (species, GMMs, and metabolites), with colours indicating variable type. Clusters of points reflect groups of variables that are positively correlated with one another, whereas variables positioned on opposite sides of the plot indicate negative correlations. (D) Clustered Image Map (a heatmap) for the variables selected by multiblock sPLS-DA on component 1. Euclidean distance and Complete linkage methods were used for clustering. Individuals (samples) are in columns (indicated by color by the intervention type) and selected features are in rows (indicated by color by the omic type – taxonomic (Species), functional (GMM) and metabolite (Metabolome) features. Color key – the values refer to the number of standard deviations away from the mean of a feature. By default, these are “trimmed” to within –3/+3 standard deviations

### 3.3. Univariate analysis of biodiversity and primary outcomes – the effect of intervention

Alpha-diversity metrics changes did not differ by intervention. Likewise, within-subject Bray-Curtis distances from Visit 2 (V2) to Visit 5 (V5) did not differ between interventions (Fig. 2, panel A). Longitudinal analysis of features between V2 and V5 showed that several bacterial species, functional modules and metabolites exhibited different trajectories between the PRO and PLA groups. Notably, *Faecalibacterium prausnitzii*, *Clostridiales* sp. and *Bacteroides fragilis* (Fig. 2, panel B) tended to increase over time in the probiotic arm, whereas they remained more stable in the placebo arm. Although the visit x treatment interaction terms were significant (P=0.007, P=0.007, P=0.008 for *Faecalibacterium prausnitzii*, *Clostridiales* sp. and *Bacteroides fragilis,* respectively, FDR-adjusted P values (Q) were non-significant (all Q = 0.578). These patterns should be interpreted as trends rather than definitive effects. At the functional level, although, GMMs related to glycolysis (MF0066) and arginine degradation (MF0055) showed modest time-dependent shifts between visits varied by intervention (P = 0.035 and P = 0.039, respectively, all Q = 0.719, Fig. 2, panel C), these signals were likely driven by baseline (V2) differences rather than true intervention effect. At the metabolomic level, 4-uanidinobutanoic acid (P=0.008, Q=0.090), hyodeoxycholic acid (P=0.029, Q=0.136) and L-glutamine (P=0.034, Q=0.136) all displayed greater increases in the PRO group between V2 and V5 than in the PLA group (Fig. 2, panel D), although the effect for L-glutamine was also partly driven by baseline (V2) differences. Overall, the univariate analyses suggest that the probiotic intervention induced coordinated but moderate changes in selected bacterial taxa, functional pathways and metabolites.

**Fig. 2.**
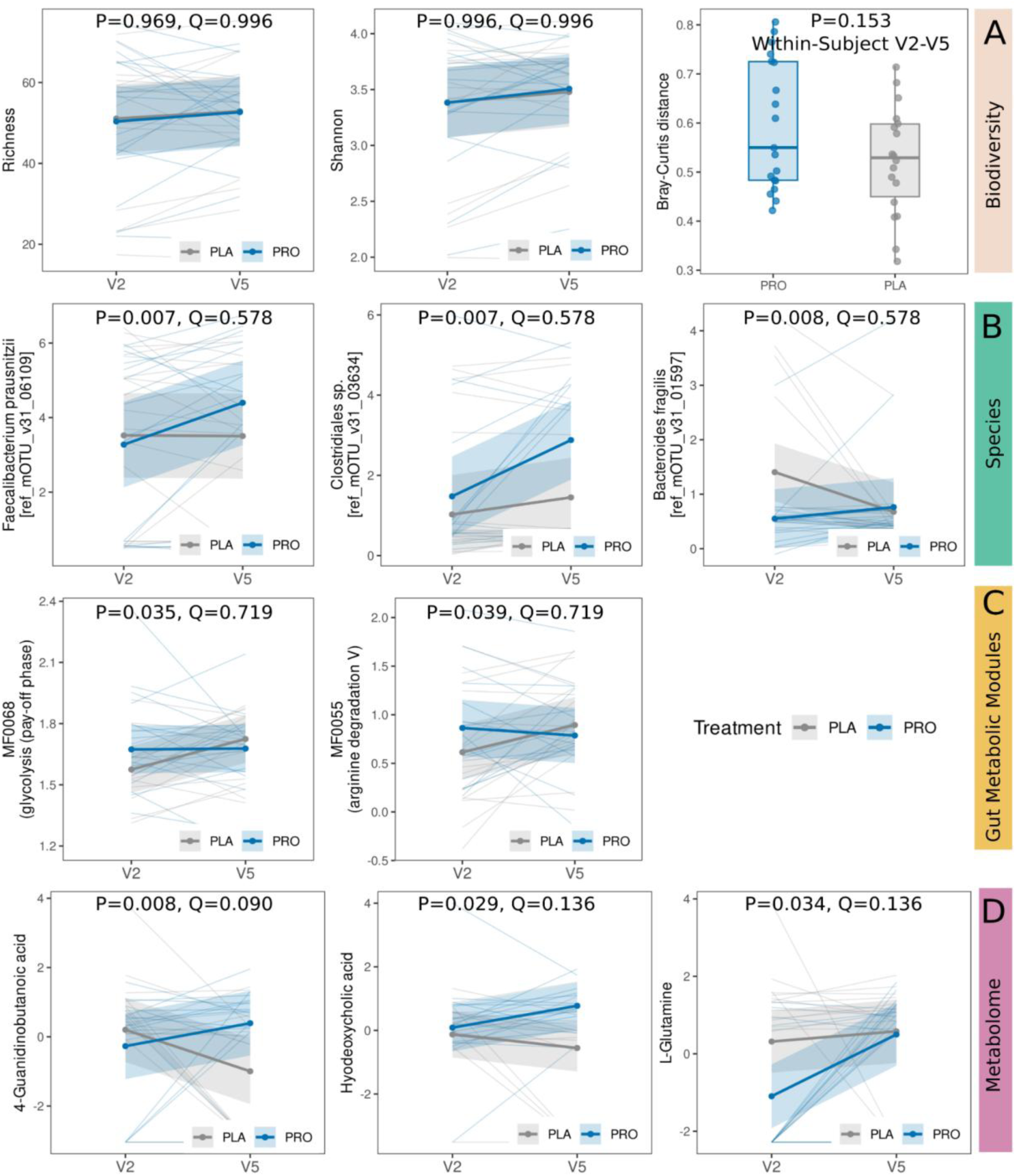
Longitudinal profiles of microbial species, gut metabolic modules (GMMs) and metabolites between Visits 2 and 5 in the probiotic and placebo groups. (A, left) Richness, (A, middle) Shannon diversity, (A, right) Within-subject V2-V5 Bray-Curtis distances comparing the PRO and PLA groups, (B) Selected bacterial species, (C) Gut Metabolic Modules (GMMs), (D) Gut metabolites; Except for the boxplot of within-subject V2-V5 Bray-Curtis distances comparing the PRO and PLA groups (A, right), all other plots represent predictor effect plots generated using the ggpredict function (R package *ggeffects*), overlaid with individual participant trajectories (“spaghetti” lines). These plots illustrate temporal changes from Visit 2 (V2) to Visit 5 (V5) in the probiotic (PRO) and placebo (PLA) groups. V2: baseline; V5: week 12

To explore the potential functional relevance of gut metabolites, we examined their associations with GI symptom burden, using both QACSMI and the simplified GI score, and treated adverse events as either numeric or binary outcomes (see Methods). In both approaches, mainly hyodeoxycholic acid showed the strongest association with a reduction in a GI score at V5 (P=0.038, Q=0.460; P=0.038, Q=0.407, for the numeric and binary outcomes, respectively, Fig. 3, Supplementary Table 1). With increasing hyodeoxycholic acid concentrations at V5, total GI score in the PRO group exhibited a decreasing trend, whereas in the PLA arm the scores remained similar across low, medium and high metabolite levels, indicating no clear dose-response relationship (Fig. 3A). Similarly, when GI AE was treated as a binary response, the PRO group revealed a decreasing probability of a total GI score above the median, while in the PLA group the pattern was reversed (Fig. 3B). Very similar patterns were observed for QACSMI, although with weaker statistical support (P=0.063, Q=0.752; P=0.093, Q=0.287, for the numeric and binary outcomes, respectively, Fig. 3C-D).

**Fig. 3.**
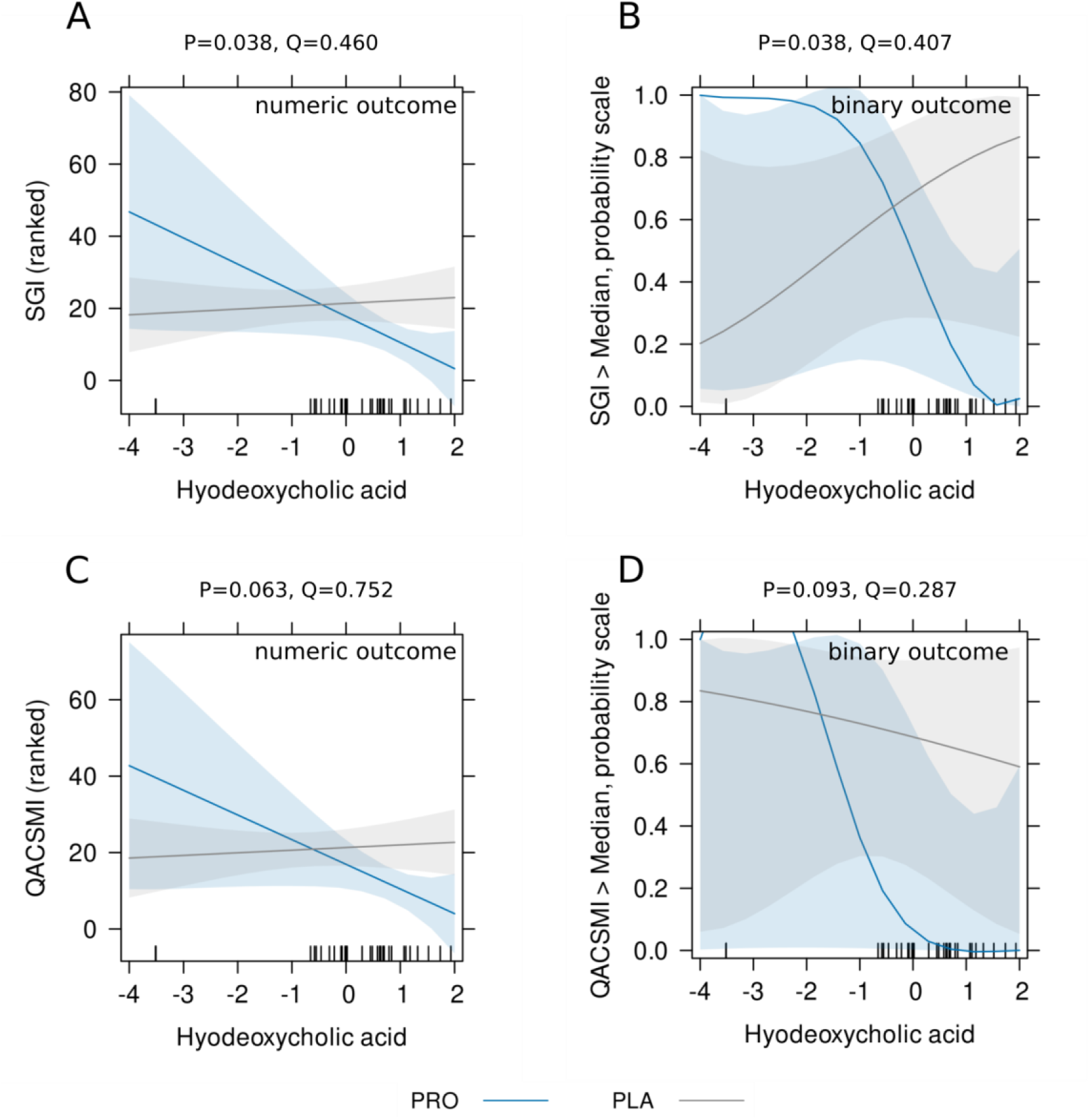
Relationship between faecal hyodeoxycholic acid levels and gastrointestinal symptom scores by treatment group. Predicted effects from linear and logistic regression models illustrating the relationship between hyodeoxycholic acid concentrations and GI symptom burden at V5. (A) Effect plot showing the fitted association between hyodeoxycholic acid and the continuous total GI score in the PRO and PLA groups, with 95% confidence bands. (B) Logistic effect plot showing the predicted probability of exhibiting a total GI score above the median (7) as a function of hyodeoxycholic acid, stratified by treatment group. (C-D) Corresponding effect plots for the QACSMI score, presented for both the numeric outcome (C) and the binary outcome, median 10 (D). Shaded regions represent 95% confidence intervals. Rugs displayed along the x-axis indicate the distribution of observed hyodeoxycholic acid concentrations in the underlying dataset (short tick marks showing individual data points). Predicted values were generated using the effects package in R.

## 4. Discussion

Our findings extend the clinical outcomes of the ProGasMet trial [15] by suggesting, for the first time, that probiotic supplementation in metformin-intolerant T2DM individuals may be associated with coordinated shifts across omic layers, particularly involving bile acid-related metabolites in particular. In the context of modulation of gut microbiota by metformin, our observation of coordinated taxonomic-functional-metabolic shifts, with bile acid-related metabolites as prominent contributors, provides a mechanistically coherent extension of the clinical ProGasMet findings.

Using a multi-block partial least squares framework (DIABLO) to jointly model species-level taxonomic profiles, gut metabolic modules (GMMs), and faecal metabolomic data, we observed moderate correlations (r = 0.4-0.6) among the dominant latent components of the three omics layers. This level of concordance suggests that taxonomic, functional, and metabolic shifts are not occurring in isolation, but rather align along a shared axis of intervention-associated change. While the treatment groups showed partial overlap, reflecting likely considerable inter-individual heterogeneity, samples tended to cluster according to intervention status at Visit 5 (12 week) across all omics layers. This consistent group differentiation implies that probiotic supplementation induces a discernible, multi-dimensional signature that transcends baseline variability and is detectable despite the modest sample size. Within the metabolomics layer, bile acid-related compounds, including hyodeoxycholic acid, chenodeoxycholic acid, and 4-guanidinobutanoic acid, were among the strongest drivers of separation and were generally enriched in the probiotic group. In contrast, metabolites such as nutriacholic acid and oleamide were more characteristic of placebo-treated individuals. When taxonomic, functional, and metabolic features selected by the model were jointly visualised, probiotic and placebo samples formed three distinct clusters, each defined by different combinations of commensal taxa, metabolic modules, and metabolites (e.g. bile acid-related metabolites), with differing profiles in the PRO-enriched and PLA-enriched groups. At the same time, the observed partial overlap between groups and the contribution of many features with small-to-moderate effects underscore the complexity of the probiotic response [51]. These results indicate that the effect is not attributable to a single taxon or metabolite. Instead, they support a systems interpretation in which probiotics induce coordinated, multifaceted changes in gut ecology and host-microbe interactions.

Complementary feature-level analyses yielded largely similar findings to those observed in the exploratory multi-omics approach. Especially in the metabolomics domain, where levels of 4-guanidinobutanoic acid, hyodeoxycholic acid, and L-glutamine increased more substantially over time in the probiotic group compared to placebo (P = 0.008, 0.029, and 0.034; Q = 0.090, 0.124, and 0.124, respectively), though the pattern for L-glutamine also reflected an initial baseline difference. These metabolite changes are consistent with broader studies demonstrating that both metformin and probiotics can remodel intestinal bile acid pools and other microbial metabolites in ways that impact host physiology. Metformin has been shown to alter gut microbiota composition and bile acid handling, including increased levels of *Akkermansia* and *Escherichia* and changes in lipopolysaccharide and SCFA production [3,5,8]. Multi-species probiotic can, in turn, promote bile salt hydrolase activity and 7α-dehydroxylation, leading to increased formation of secondary bile acids such as deoxycholic acid, chenodeoxycholic acid and hyocholic acid, and can shift the microbiota away from gas– and toxin-producing taxa [32,52]. Importantly, these patterns in our dataset emerged after adjusting for metformin dose as a time-varying covariate in our longitudinal models, indicating that the observed differences are unlikely to be driven by variation in metformin exposure.

Bile acids occupy a central position at the interface between the gut microbiota, host metabolism and metformin’s gut-mediated actions. Beyond their classical role, they are now recognized as signaling molecules that regulate their own synthesis and transport, as well as triglyceride, cholesterol and glucose homeostasis via FXR– and TGR5-dependent pathways [24,28,53,54]. In enteroendocrine L-cells, FXR activation generally restrains GLP-1 production, whereas TGR5 activation stimulates GLP-1 secretion via cAMP-dependent mechanisms. Metformin itself reduces active ileal bile acid reabsorption through ASBT inhibition, thereby increasing luminal bile acid exposure and enhancing GLP-1 secretion from L-cells, which may partly mediate its glucose-lowering effects [28,53]. At the same time, clinical and experimental data indicate that metformin treatment is frequently accompanied by bile acid disturbances, reduced reabsorption and osmotic diarrhoea in a substantial proportion of patients [13,55,56], suggesting that bile acid dysregulation may also contribute to gastrointestinal side effects.

Within this framework, our multi-omics results are notable in that bile acid–related metabolites, including hyodeoxycholic acid (HDCA), chenodeoxycholic acid and 4-guanidinobutanoic acid, emerged among the strongest contributors to the multivariate separation between probiotic and placebo arms, and were generally enriched in the probiotic group. When we explicitly linked metabolites to clinical outcomes, HDCA repeatedly appeared as the compound most strongly associated with reduced gastrointestinal symptom burden in probiotic-treated patients. Using both the Metformin Symptom Severity Score (QACSMI) and a simplified GI symptom score, and modelling adverse events as either continuous or binary outcomes, higher HDCA concentrations at Visit 5 were consistently related to lower symptom burden in the probiotic arm (numeric GI score: P = 0.038, Q = 0.460; binary outcome: P = 0.038, Q = 0.407), with similar but weaker trends for QACSMI (P = 0.063, Q = 0.752; P = 0.093, Q = 0.287). Importantly, the shape of this relationship differed by treatment group: in probiotic-treated participants, GI scores declined across HDCA strata, whereas in placebo-treated individuals scores remained largely unchanged across the same concentration range. This pattern suggests that HDCA is not simply a generic marker of lower symptom burden, but that its functional coupling to GI outcomes may be context-dependent and more pronounced in the presence of probiotic supplementation.

These observations are biologically plausible in light of emerging data on hyocholic and hyodeoxycholic acid species. Hyocholic acid-type bile acids have been identified as regulators of glucose homeostasis acting through a distinct pattern of TGR5 and FXR signalling, improving metabolic parameters in experimental models [57]. HDCA itself has been shown to alleviate hyperlipidaemia, improve glucose metabolism and ameliorate metabolic associated steatotic liver disease (MASLD) in animal studies [57–60]. More broadly, secondary bile acids such as deoxycholic acid and lithocholic acid can exert anti-inflammatory effects, modulate innate and adaptive immunity and influence glucagon-like peptide-1 (GLP-1) secretion via TGR5 activation [26,61–63]. Probiotic interventions, including multi-species complexes and specific strains such as *Bacillus subtilis*, have been reported to remodel bile acid pools, promote the conversion of primary to secondary bile acids and shift metabolic and inflammatory profiles in models of T2DM and liver disease [32,52,55]. Beyond its metabolic signalling properties, HDCA has also been proposed as a bile acid excipient to optimise metformin delivery: in preclinical work, an HDCA:metformin formulation outperformed unmodified metformin in lowering fasting glucose, increasing GLP-1, improving oxidative stress markers and reducing metformin-related side effects [64].

Taken together, these mechanistic insights and our exploratory findings support the hypothesis that, in metformin-intolerant individuals, multi-strain probiotic supplementation may promote a bile acid configuration in which higher HDCA levels are associated with better GI tolerance. One plausible interpretation is that probiotic-induced changes in the microbiota enhance the conversion of primary to secondary bile acids in a way that preserves metabolic benefits while mitigating metformin-related diarrhoea and bloating, for example, *via* more favourable FXR/TGR5 signalling, reduced intestinal inflammation, improved barrier function or altered motility [28,55,65]. While the present results are exploratory and did not withstand stringent multiple-testing correction, the internal consistency across methods and the biological plausibility of HDCA as a mediator make this axis a compelling target for confirmation in larger, mechanistically focused studies. Against this bile acid-centred background, it is noteworthy that probiotic supplementation did not remodel the microbiome at the level of global biodiversity. Alpha-diversity metrics remained stable across timepoints and did not differ meaningfully between treatment arms. Similarly, within-subject Bray-Curtis distances from Visit 2 to Visit 5 were comparable between the probiotic and placebo groups, suggesting no significant differences in overall richness, evenness, or community-wide turnover. Supporting this interpretation, in our longitudinal univariate analyses, probiotic supplementation was associated with coordinated, though modest, shifts in selected bacterial taxa, functional modules, and metabolites, rather than with broad-scale restructuring of the gut microbiome. Over the 12-week period from Visit 2 to Visit 5, several features exhibited differential trajectories between the probiotic and placebo arms. Canonically beneficial taxa such as *Faecalibacterium prausnitzii*, a *Clostridiales* species, and *Bacteroides fragilis* tended to increase in relative abundance among probiotic-treated participants, while remaining comparatively stable in the placebo group. The corresponding visit × treatment interaction terms reached nominal significance (P = 0.007, 0.007, and 0.008, respectively), but did not remain significant after correction for multiple testing (Q = 0.578 for all). These findings are therefore best interpreted as indicative trends consistent with a probiotic effect, rather than as definitive evidence of taxon-level change. Nevertheless, the directionality is notable, given that *F. prausnitzii* and related butyrate-producing taxa are consistently reported as depleted in T2DM and associated with an increased inflammatory tone in the gut [19,20]. Even modest increases in such commensals could therefore be biologically meaningful in the context of metformin intolerance.

At the functional level, gut metabolic modules involved in glycolysis (MF0066) and arginine degradation (MF0055) also demonstrated modest time-dependent changes that diverged between treatment arms (P = 0.035 and 0.039; Q = 0.719 for both). However, these effects appear at least partially attributable to baseline imbalances at Visit 2 rather than clear probiotic-induced shifts. These findings suggest a coherent directional effect of probiotic treatment, stimulating the gut ecosystem toward increased levels of several microbial and metabolic features that may support anti-inflammatory activity or epithelial barrier integrity. However, the modest effect sizes, potential influence of baseline differences, and sensitivity of most associations to multiple-testing correction emphasize the exploratory nature of these findings. Although the magnitude of these changes is small and sensitivity to multiple-testing adjustments, the repeated involvement of biologically plausible anti-inflammatory taxa and bile acid-related metabolites presents compelling candidates that may underlie the improved metformin tolerance observed in the parent trial.

### 4.1. Strengths

From a mechanistic standpoint, the study’s strength lies in its truly integrative, multi-omics approach. The application of multi-block partial least squares modelling, alongside covariate-adjusted univariate tests, yields converging lines of evidence linking specific taxa and metabolites, most notably bile acids such as HDCA, to GI symptom outcomes. Importantly, this analysis was restricted to the first treatment period, prior to any crossover-related effects that might confound the microbiome. This methodological choice avoids the carry-over signal observed clinically in participants who switched from probiotic to placebo and allows for a clean parallel-group comparison of probiotic versus placebo effects.

Although modest in size, the cohort is clinically well characterized. All participants had T2DM, obesity, and metformin GI intolerance, precisely the population for whom alternative strategies to improve drug tolerability are most relevant. The availability of paired stool samples at baseline and after 12 weeks, along with rich phenotypic and clinical data, provides a valuable resource for dissecting microbiota-drug-symptom interactions in a real-world, high-need population.

### 4.2. Limitations

The principal limitation of this study is its modest sample size. Only 34 participants had complete multi-omics data from the first treatment period, which substantially limits statistical power, particularly in the context of high-dimensional microbiome and metabolomics analyses. The limitation of the multiomics analysis to only the first period of the cross-over study was due to the presence of a significant carry-over effect between the study periods, which was observed in the case of clinical symptoms [15] Because standard two-period cross-over inference relies on the assumption of no carry-over effects [66], and because intervention-related shifts in the gut microbiota and metabolome may plausibly persist beyond short washout intervals [67], limiting analyses to period 1 provides the most conservative and interpretable dataset for attributing observed multi-omics differences to the assigned intervention. This constraint likely accounts for the limited number of associations that survived correction for multiple testing, despite the presence of moderate effect sizes and consistent patterns across related taxa and metabolites. Consequently, the exploratory nature of our findings, and the emphasis on nominal P values and effect directions to highlight candidates such as *Collinsella*-related taxa and HDCA, necessitates cautious interpretation and underscores the need for independent replication.

Moreover, our associations between microbial features, bile acids, and symptom scores remain observational in nature and cannot establish causality. It is unclear whether these associations reflect direct mechanistic effects, secondary consequences of symptom severity, or shared upstream determinants.

The study population, although clinically well-defined with respect to diagnosis and metformin intolerance, was recruited from a single centre and comprised predominantly older, obese individuals with limited ethnic and geographic diversity. This limits the generalisability of our findings to younger populations, individuals with differing BMI profiles, other ethnic backgrounds, or those treated outside a structured metformin titration protocol. Furthermore, only a single probiotic formulation and dosing schedule was evaluated. Thus, the observed multi-omics signatures may not extend to other strains, dosages, or combinations.

Finally, although we included a broad untargeted stool metabolomics panel, encompassing bile acids, and inferred functional capacity from taxonomic data, we did not directly assess host-level endpoints such as mucosal gene expression, bile acid receptor activation, or epithelial barrier integrity. As such, the proposed mechanistic links between probiotic-induced microbial changes, altered bile acid profiles, and GI symptoms warrant experimental validation.

Although our study has some abovementioned limitations, it is worth highliting its novelty and clinical meaning.

## 5. Conclusions

Hyodeoxycholic acid emerged as a noteworthy candidate linking multi-strain probiotic supplementation to reduce metformin-associated AEs. Although some associations did not remain significant after multiple testing correction, their coherence and biological plausibility suggest that probiotic-induced remodeling of bile acid metabolism, alongside shifts in microbial taxa, may contribute to improved metformin tolerance. These findings show exploratory links to GI symptom burden and offer a foundation for future mechanistic studies on a larger scale. Collectively, the results support the broader concept that microbiome-targeted interventions may offer a promising strategy to mitigate drug-induced GI intolerance in diabetes care. Moreover, the observed increase in HDCA suggests that such probiotic formulation may have potential benefits beyond metformin intolerance, particularly in applications where modulation of bile acid profiles is therapeutically relevant.

## Funding

This study was supported by 2020 research grant from the Diabetes Poland as a research project entitled “Diabetes Poland Competition for scientific grant” and by statutory finances of Medical University of Silesia (PCN-1–176/N/9/K). The probiotics were donated by a distributor of probiotic formulas Sanprobi sp. z o. o. sp. k. The funding sources had no role in the study design, data collection, interpretation of analysis, writing of the manuscript, or decision to submit the publication.

**Data availability –** raw data and metadata are available under accession number PRJNA1378533

**CRediT authorship contribution statement** (Conceptualization, Data curation, Formal analysis, Funding acquisition, Investigation, Methodology, Project administration, Resources, Software, Supervision, Validation, Visualization, Writing – original draft, Writing – review and editing)

**Hanna Kwiendacz**: Conceptualization, Project administration, Resources, Writing – original draft **Danuta Cembrowska-Lech**: Data curation, Formal analysis, Software, Visualization, Writing – original draft, **Karolina Skonieczna-Żydecka**: Conceptualization, Investigation, Methodology, Resources **Karolina Klimontowicz**: Investigation, Methodology, Resources, **Konrad Podsiadło**: Data curation, Formal analysis, Visualization, Writing – original draft, **Anna Wierzbicka-Woś**: Investigation, Methodology **Daniel Styburski**: Investigation, Methodology **Mariusz Kaczmarczyk**: Data curation, Formal analysis, Visualization, Writing – original draft **Janusz Gumprecht**: Supervision, Validation, Writing – original draft, **Igor Łoniewski**: Conceptualization, Resources, Supervision, Project administration, Writing – original draft **Katarzyna Nabrdalik**: Conceptualization, Resources, Supervision, Project administration, Writing – original draft

## Data Availability

Raw data and metadata are available under accession number PRJNA1378533

https://www.ncbi.nlm.nih.gov/bioproject/1378533

