## Supplementary Materials for "Multi-strain probiotic enhances metformin tolerance by modulating gut microbiome and bile acid pathways: Insight from multi-omics post-hoc analysis (ProGasMet trial)"

**Supplementary Figure 1.** Classification error rates (overall and balanced) with respect to the number of components for three distances

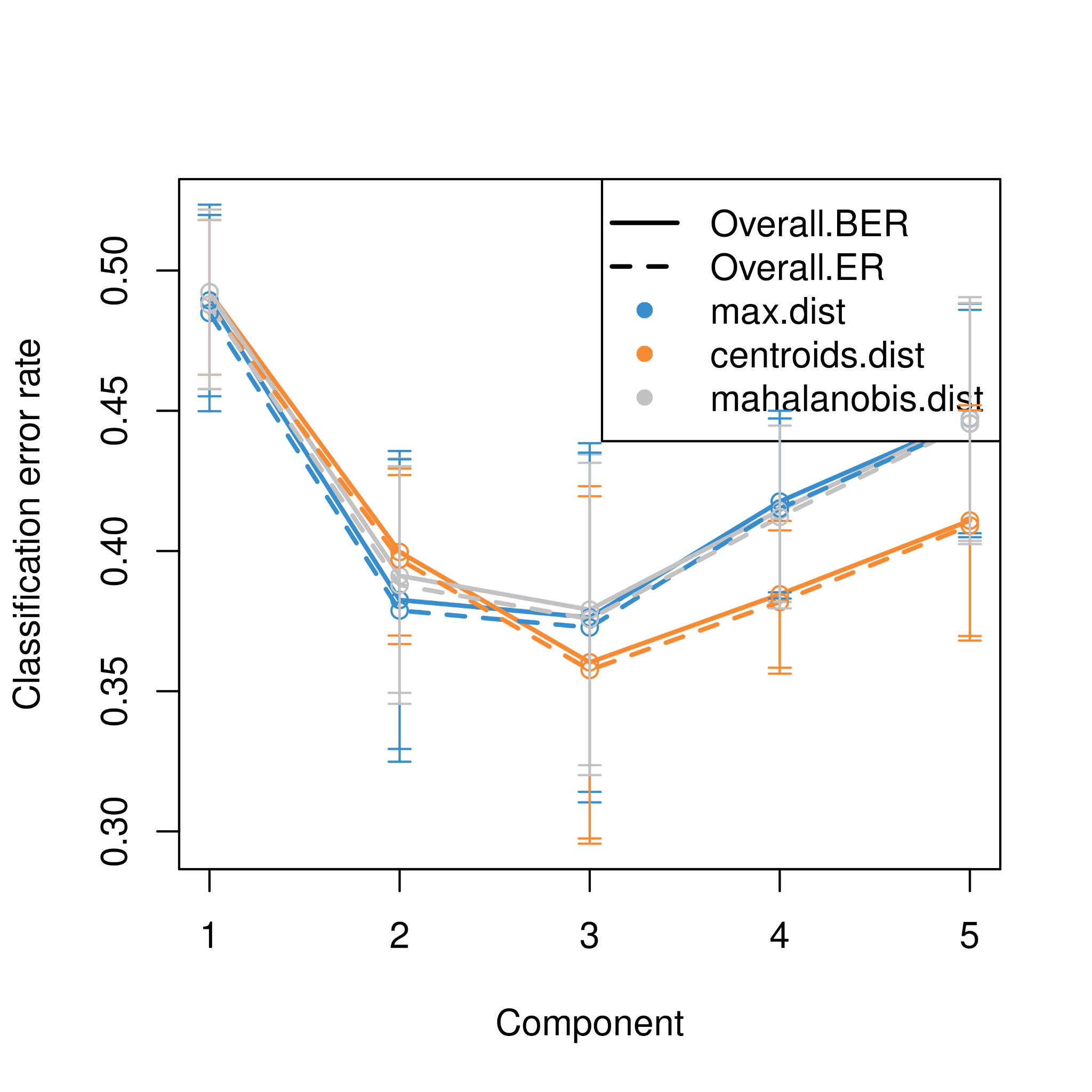

N-integration with Projection to Latent Structures models (PLS) with Discriminant Analysis (PLS-DA) using 10-fold cross validation with 10 repeats. Y-axis - classification error rates: overall - Overall.ER and balanced - Overall.BER. X-axis - the number of components for three different distances. Bars represent the standard deviation from the 10 repeated folds.

The minimum error rate occurs for 3 components and centroid distances (centroids.dist).

**Supplementary Figure 2. Feature importance for each of the block**

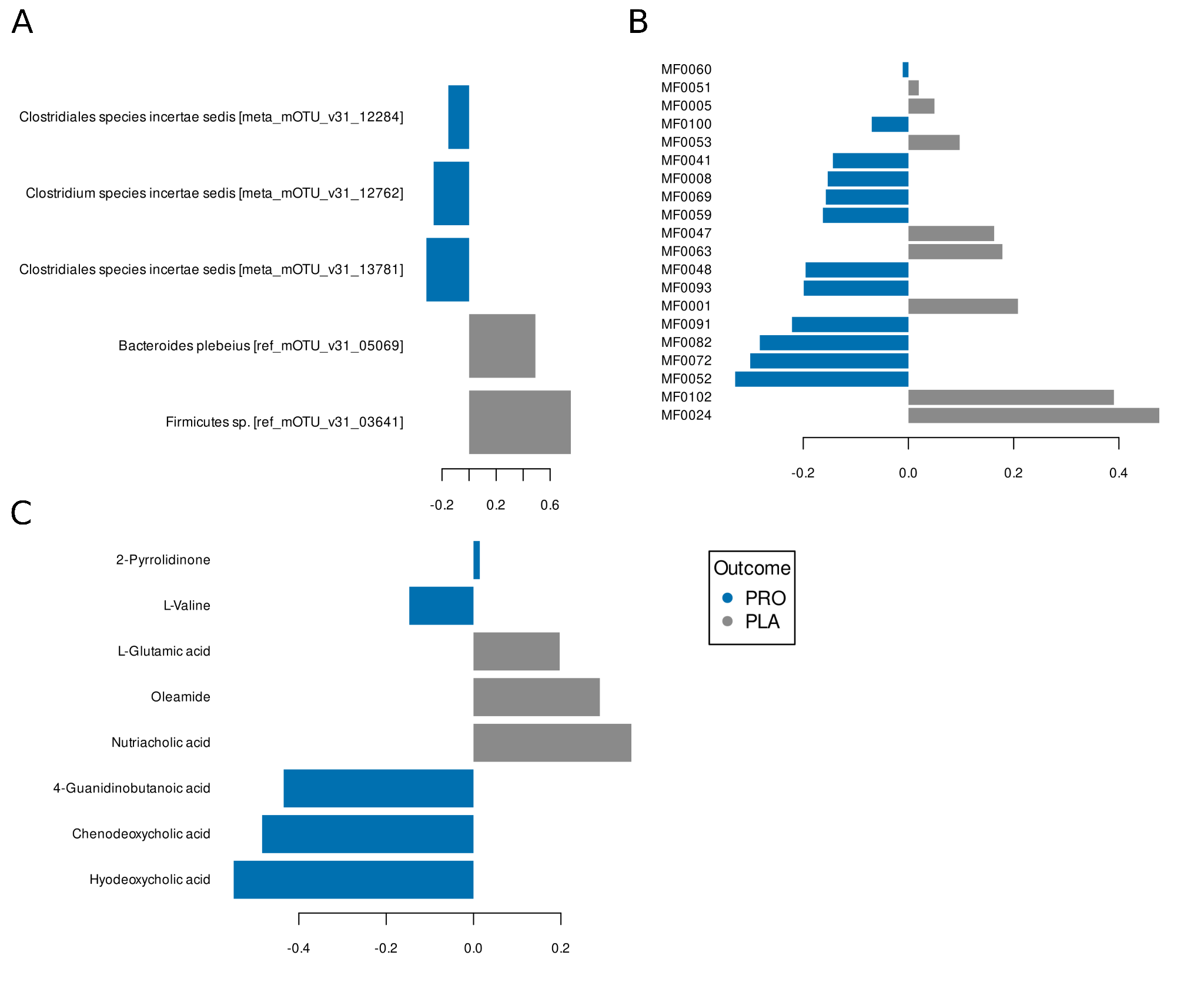

A - Taxonomic block (species level), B - Functional block (Gut Metabolic Modules), C - Metabolomic block

The loading plot shows the multi-omics signature selected on component 1, with each panel representing a different data type. The contribution of each variable is visualised by the length of the bar (i.e. the absolute value of its loading coefficient). The selected features are associated with one of the two interventions (PRO - probiotic or PLA - placebo).
